# Patient Registry of Upper Limb Outcomes (PRULO) A Protocol for an orthopaedic clinical quality registry to monitor treatment outcomes

**DOI:** 10.1101/2023.02.01.23284494

**Authors:** Corey Scholes, Kevin Eng, Meredith Harrison-Brown, Milad Ebrahimi, Graeme Brown, Stephen Gill, Richard Page

**Affiliations:** EBM Analytics; Barwon Centre for Orthopaedic Research and Education (B-CORE); Geelong Orthopaedics; School of Medicine, Deakin University

## Abstract

**Introduction:** Registries have been widely utilised to track clinical results in observational cohorts for quality improvement. They have been very successful in orthopaedics, particularly in the context of arthroplasties where they have guided and optimised implant choice, patient safety and identified underperforming implants. However, equivalent systems to monitor outcomes in soft-tissue disorders are lacking. This manuscript describes the protocols around design, ethics and implementation of a regional registry focusing on upper limb soft tissue orthopaedic conditions.

**Methods and analysis:** PRULO (Patient Registry of Upper Limb pathology Outcomes) is a multi-cohort, prospective observational, clinical quality registry collating clinical data and patient-reported outcomes for patients presenting to a specialist orthopaedic clinic with upper limb pathology. PRULO is currently a single-centre study involving three clinician investigators, which aims to determine what patient characteristics, pathology factors and treatment strategies are associated with treatment success within 2 years of surgical or non-surgical treatment of targeted pathology. PRULO captures patient reported outcomes (VAS, EQ5D-5L, QuickDASH, MODEMS-Expectations and Satisfaction, WORC, WOSI), clinical and radiological data. Data points are recorded at initial practice registration, after initial consultation, intraoperatively, as well as 3, 6, 12 and 24 months. Inclusion criteria are patients aged 16 and above offered treatment by speciality trained surgeons, for upper limb orthopaedic pathology. Patient subgroups (cohorts) will include shoulder conditions: conditions affecting predominantly the rotator cuff (tear, tendinopathy), conditions associated with glenohumeral instability, as well as all other conditions presenting in the shoulder elbow, hand and wrist, according to the surgeon-generated diagnosis.

**Ethics and dissemination:** Ethical approval was obtained by the Barwon Health Research Ethics Committee (approval number 19/70).

**Trial Registration:** The project was added as a trial to the Australia and New Zealand Clinical Trials Registry (ACTRN12619000770167) 23-May-2019.

## INTRODUCTION

### Clinical Background

Clinical quality registries are defined as “an organised system that uses observational study methods to collect uniform data (clinical and other) to evaluate specified outcomes for a population defined by a particular disease, condition or exposure, that serves a predetermined scientific, clinical or policy purpose(s)” (Gliklich et al., 2014). Registries contribute evidence to optimise patient outcomes. In orthopaedics, many examples of national clinical registries exist such as the Australian Orthopaedic Association National Joint Replacement Registry and other registries in Sweden, England and New Zealand (de Steiger et al., 2013; Hooper et al., 2014; Kärrholm, 2010; Porter et al., 2019). By tracking multiple data points over time, these registries have been highly successful in providing population based evidence for identifying underperforming prosthesis and surgeons, saving many millions of dollars in low value surgeries, reducing the burden of revision surgery and ultimately improving patient care in a timely manner (Gill & Page, 2019). Registries require extensive planning, implementation, cost and analysis, and these can fall outside the capacity of smaller surgical practices and hospitals (Colwell et al., 2009). Soft tissue shoulder procedures have traditionally not been included in national orthopaedic registries.

#### Regional registry for upper limb pathology

The clinician-investigators for the current registry consult at a private specialist orthopaedic clinic, which provides the main private orthopaedic services in the region. Surgical facilities in the region consist of one government funded public hospital and two private hospitals. The clinic has been providing orthopaedic services since 1996 and has gradually expanded and sub-specialised as the population has grown. The investigators contribute shoulder arthroplasty data to national and hospital-based regional registries. Results for orthopaedic surgeries not covered by arthroplasty registries have traditionally been collected by individual surgeons on paper or commercially available electronic databases without an overarching data governance model. This approach has been limited by poor standardisation across clinics. Without a standardised approach to collecting outcomes for non-arthroplasty patients, consolidation of the data into a single dataset for subsequent analysis has been historically difficult. In 2013, an attempt was made to capture patient data using a commercial database and internet based patient reported outcome measures (PROMS) using an electronic tablet in the clinic with reminders via email. This process was successfully implemented into the investigators’ practice systems, however fewer than 20% of patients responded to follow up after their initial surgeries. It was recognised that the clinic had technological capability, but lacked the resources to ensure data quality and achieve consistently high patient follow up.

### Objectives

The primary purpose of the registry is to operate as a technical platform to monitor patient outcomes associated with orthopaedic upper limb pathology, initially focussing on shoulder rotator cuff repair and shoulder instability surgery within a multi-surgeon private practice based in regional Australia. It will capture and organise clinical, functional, imaging and patient-reported data, enabling temporal and cross sectional comparisons within and between pathological groups (cohorts). It is titled the Patient Registry of Upper Limb pathology Outcomes (PRULO). The ultimate vision of PRULO is to expand to a broad group of upper limb orthopaedic subspecialty clinics in Australia. The objective of this protocol is to provide the rationale, methodological descriptions and technical tools used to establish and maintain the registry. By publishing this protocol the investigators hope to provide an example of how this can be done on scale to suit any practice, to help overcome impediments that might be identified early, and to enable broader data capture.

## METHODS

### Design and setting

PRULO is a prospective observational clinical quality registry that collates clinical data and patient outcomes for patients presenting to a private clinic with orthopaedic (or musculoskeletal) upper limb pathology (Figure 1). This is a single practice registry, with three surgeon investigators consulting from multiple locations and operating in private and public hospitals. The registry is integrated into usual clinical care pathways, utilising a “standard of care” cohort design, with outcome measures selected according to presenting pathology (Figure 1). All patient data will be collected and stored together in a single registry.

**Figure 1:**
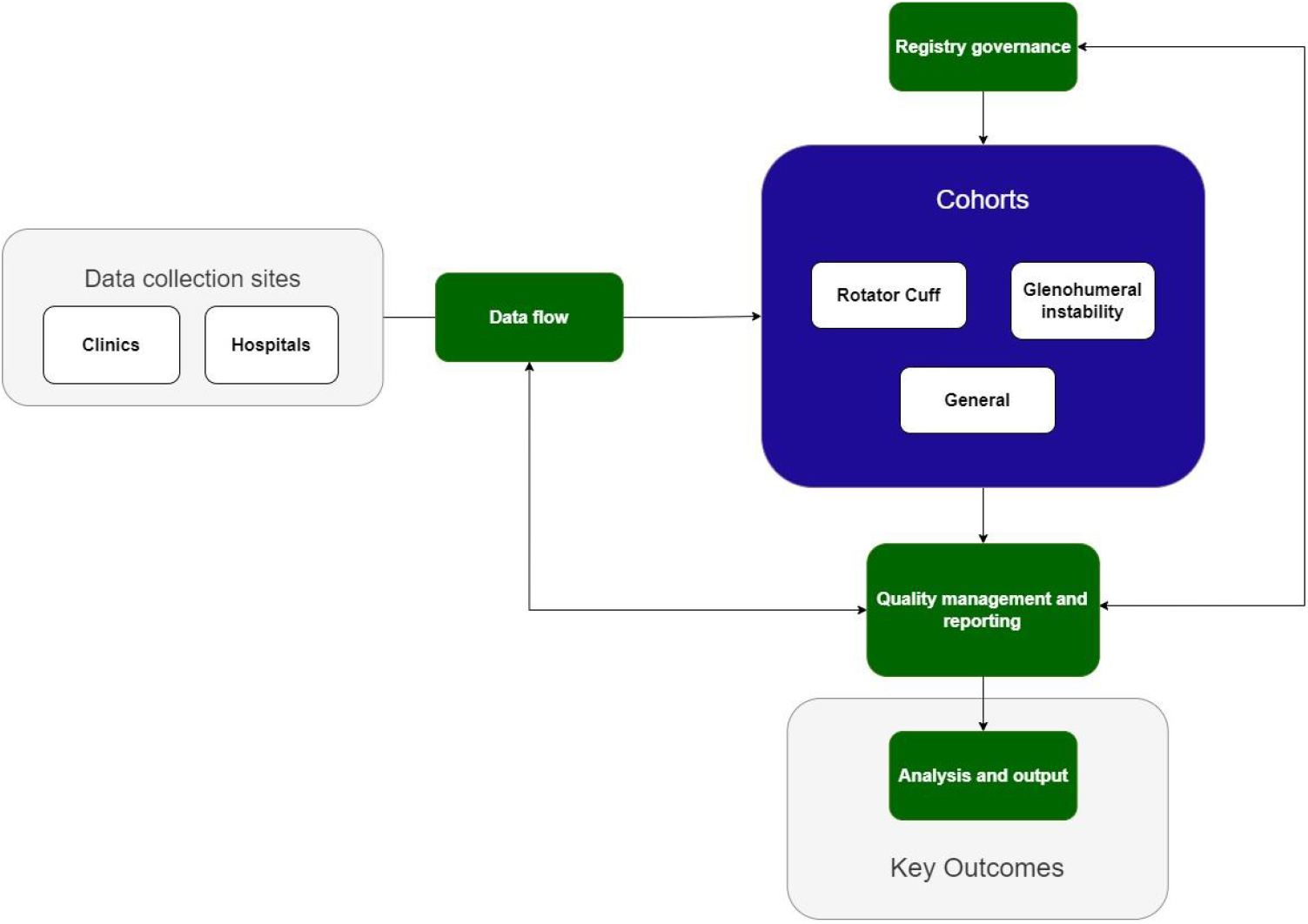
Overview of the key components of the PRULO Registry

### Process to establish PRULO registry

A design and preparation process was undertaken to establish the registry structure (including steering committee and contributing practices), governance framework and analytical plans of the component cohorts. Documentation was produced and HREC approval granted, while technical systems were in development and funding was secured to commence prospective operation of the registry.

The first patients were entered into PRULO in 2020 starting with a single contributing surgeon. The data management team conducted training on procedures with the surgeon and administrative staff via webcall. Procedures were published for staff including clinic consultation data collection, staff follow up lists, operating list data, complications and discrepancy lists for filling in missed data. All procedures have been published online to allow staff to access them anytime. The second surgeon was added approximately 3 months later and the third 3 months after that to allow focus on training staff and working with individual variations in practice structure.

### Participant Eligibility and Recruitment - Registry

#### Patient inclusion/exclusion

All patients presenting to the participating surgeons for treatment are eligible for inclusion in the registry under an opt-out consent arrangement, providing they meet the following criteria:

#### Inclusion Criteria

1. Presenting for surgical review with orthopaedic shoulder, elbow, hand or wrist pathology
  - The presentation is the first time the patient has presented any one of the participating orthopaedic surgeons
  - The presentation is a new pathology for a patient offered definitive treatment (surgical or non-surgical) for a presenting pathology of the upper limb (non-surgical treatment may comprise referral to physiotherapy; intra-articular injection; bracing or other aids)

#### Exclusion Criteria

1. Psychiatric or neurological illness that precludes consent to orthopaedic treatment
2. Under 16 years of age at consultation
3. Revocation of consent for research use of personal data
4. Subsequent aetiology of presenting pathology is deemed to be of non-upper limb origin (e.g. cervicogenic pain)

### Recruitment and Consent

Eligible patients are presented with a Participant Information Sheet and a withdrawal of consent form via email or SMS following registration with the clinic, or in person by the consultant surgeon, or front desk personnel. They are given the opportunity to ask any questions and may opt out at any time before, during or after their treatment by returning a withdrawal of consent form or by informing practice staff. Participants are also able to opt out of receiving requests to complete patient reported outcome measures and retain their existing data in the registry if they wish.

#### Recruitment progress

Participant recruitment and data collection began in October 2020. From its inception in 2020 to the 30th September 2022, 1630 patients have been recruited into the PRULO registry, with a rate of subsequent withdrawal of approximately 1% for patient-reported outcomes, and 0.3% for the registry entirely. Approximately 129 of the 1632 patients have reached the final (24 months) window for followup by the registry.

### Data sources

Participants are sourced from the participating clinic and hospital locations (Figure 2) when they present to the participating surgeons for treatment of eligible upper limb orthopaedic conditions. Clinical information is collected from patients by electronic forms. Electronic notes are recorded during the surgeon consultation as routine. Notes, imaging, pathology, and letters are inputted or transferred to the practice management system (PMS) as normal. The PMS is the primary source of data for the PRULO registry database. Patient-reported outcome measures (PROMS) are collected electronically from patients sent via SMS or email using secure electronic questionnaires linked to Australian servers (QuestionPro, Austin, TX, USA), and transferred directly into the PRULO database.

**Figure 2:**
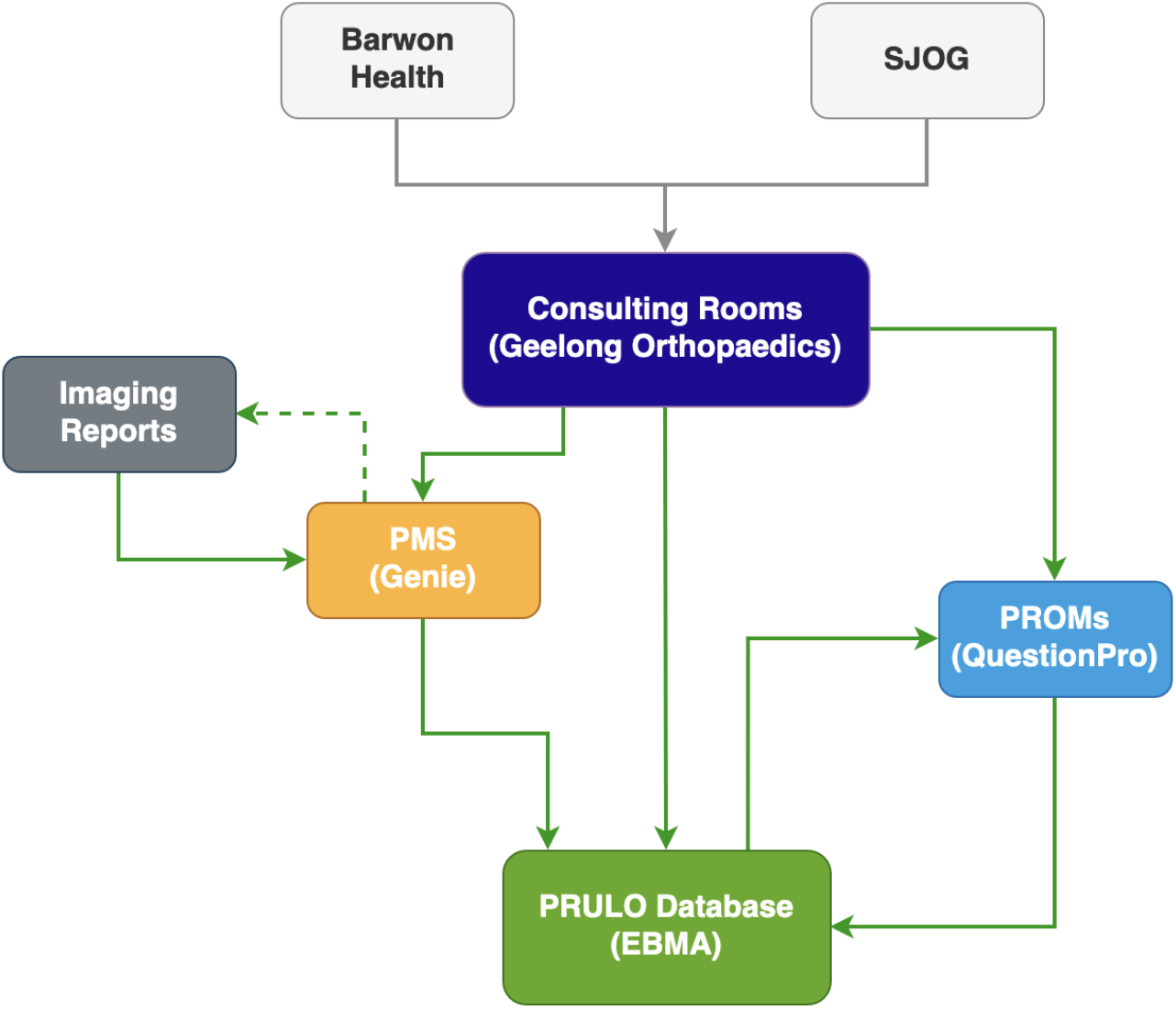
Data source flow of registry

Data collected within the practice management system (Table 1) for inclusion in the registry include:

**Table 1:**
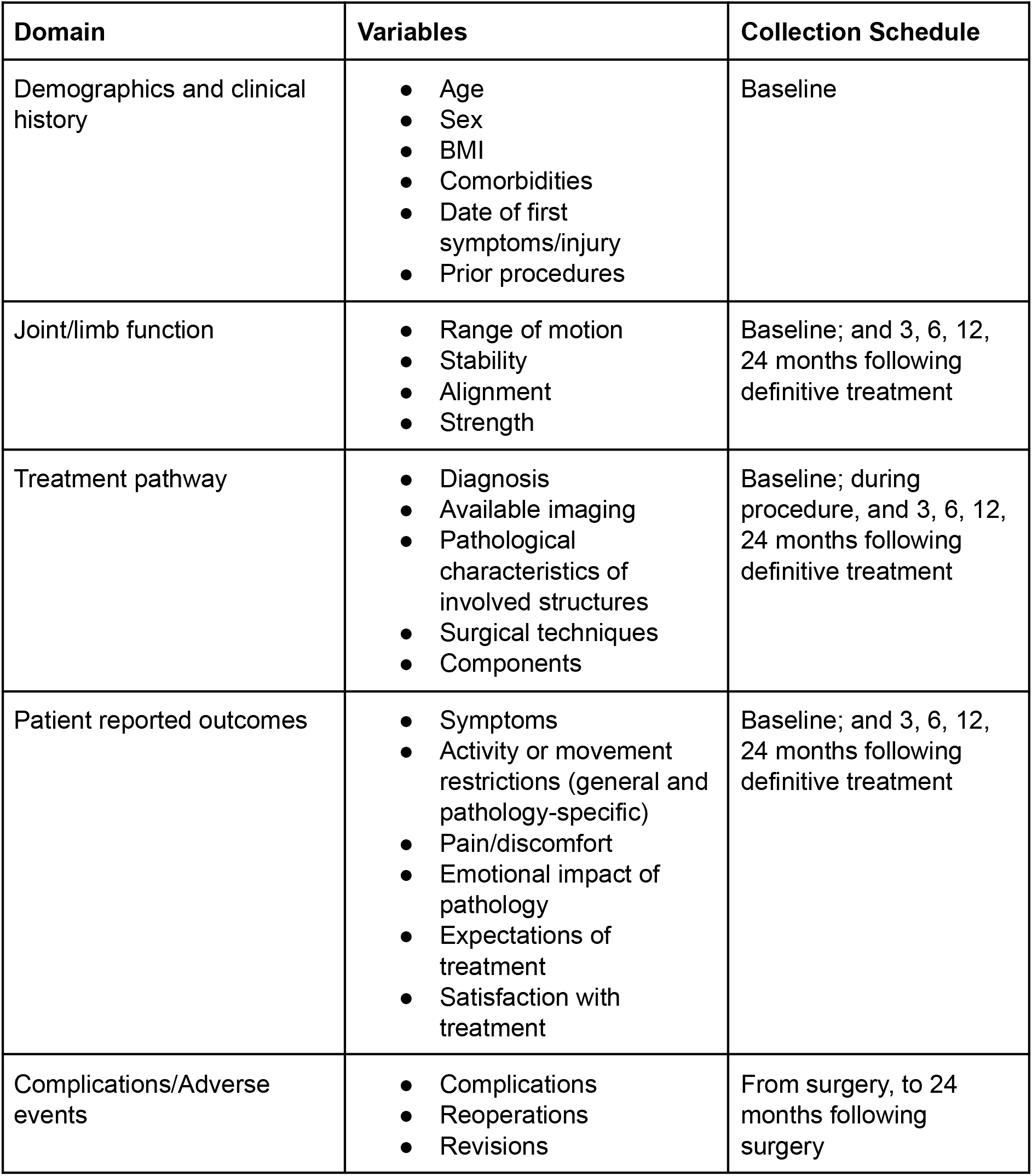
Outline of routinely collected data entered into PRULO

- Patient data: demographic data and medical history are recorded from the records made during patient consultation.
- Clinical and treatment data: radiological reports (x-ray, CT or MRI) collected routinely as part of diagnosis, surgical planning and postoperative follow up. The diagnosis, mode of treatment (non operative vs operative), surgical interventions, timing of treatments, and complications or revision procedures are recorded from electronic medical records or during patient consultation.
- Outcomes data: findings from clinical examination and assessment of range of motion, strength, joint stability and alignment are recorded from electronic medical records. Patient-reported outcome measures (PROMs) are completed by the patient as described above.

### Participant Cohort Definitions

Specific cohorts are defined for the purpose of selecting the most relevant patient-reported outcome measures, which may include general and pathology-specific questionnaires.

Specific surgical cohorts include:

- Shoulder - Rotator cuff: Primary diagnosis of rotator cuff pathology including subacromial impingement, rotator cuff tendinopathy, rotator cuff tear (intrasubstance, partial thickness, full thickness). Patients with rotator cuff pathology but not deemed to be the primary diagnosis for which the patient is seeking treatment, (for example cuff tear arthropathy) will be excluded from this cohort.
- Shoulder - Glenohumeral Instability: Diagnosis of glenohumeral instability, including dislocation with associated fracture or voluntary dislocation.
- Shoulder - General: Any shoulder pathology not covered by the above cohorts, irrespective of treatment strategy.
- Elbow pathology: Any pathology affecting the elbow will be included in this cohort.
- Wrist/hand pathology: Any pathology primarily affecting the structures of the hand and wrist will be included in this cohort.

### Data Management

Management of data collection and coordination of followup for incomplete data are managed by a data management team contracted by the participating surgeons (EBM Analytics, Sydney, NSW, Aus). The data management team processes exports from the practice management system to link registry records with appointment lists and to identify new patients eligible for recruitment. Data requirements are communicated on a daily basis by the data management team to practice staff and participating surgeons via a web-based interface for patients with an upcoming consultation or surgery. If data entry is required for one of the pre-defined registry intervals at the time of the patient visit, a unique link to the appropriate data entry form or questionnaire will be presented for communication to the surgeon or patient, as required.

#### Data cleaning

Data are assessed for quality in three domains on a routine basis;

- Completeness - does the intended field have a response; If so,
- Validity - does the field response conform with relevant rules to be considered a true and accurate response; If so,
- Consistency - is the field response compatible with other field responses for the same patient record

The cleaning process occurs in layers. The first layer occurs shortly (2-4 weeks) after a patient consultation, with particular attention paid to patients presenting for an initial consultation with the surgeon. Fields unavailable at the time of the appointment (e.g. diagnosis; affected side; treatment type offered) are captured and inserted back into the database. The second layer is to withdraw any record with clinical fields that fail to meet certain quality criteria (missing, invalid, inconsistent) which are manually addressed through chart review. Thirdly, when subsets of records are retrieved for a specific analysis, additional record checking is performed to i) confirm inclusion and exclusion criteria; ii) provide identifiers for potential data linkage; iii) retrieve additional fields specific to the analysis. Updates to individual records agreed between the investigators and the registry management team retrieved for the analysis are bulk-inserted back into the database.

### Governance

A governance and steering committee with representatives from the research investigators, data management team and IT providers will maintain the governance of PRULO. Quality assurance of the registry will be maintained through regular auditing and quarterly reporting of data completeness, consistency and validity. Audit reporting will be provided to the registry steering committee and communicated to the surgeon contributors. Regular monitoring of registry data will be maintained by the data management team to ensure security has not been breached.

#### Termination

The study will be led and funded by the principal surgeons and is intended to be ongoing. The registry may be terminated on the agreement of the investigators if it is no longer required, or if funding cannot be secured for ongoing management. The research dataset will be retained securely at Geelong Orthopaedics.

## DISCUSSION

The investigators look forward to publishing results and analysis. The design allows the registry to expand to include new surgeons who join the investigators to contribute their data. This registry will form the backbone of surgical results for non arthroplasty surgeries in the region, filling a significant gap in data, which to date focuses on arthroplasty, and providing evidence for the wider orthopaedic community.

### Limitations

#### Selection Bias

PRULO is an electronic, opt-out consent clinical registry that is vulnerable to selection bias at different stages of the data collection process collected by the registry. The selection biases inherent in PRULO are reflective of biases present in running a clinical practice in the setting described. At enrolment into the registry, patients are contacted via phone text message or by email to alert them to the presence of the registry within the practice and provide the relevant patient information. The capacity for patients to engage in this process may be limited by language, technological and competency barriers. Other sources of selection bias emerge from public patients presenting for initial review in the rooms and going on to have treatment and subsequent management within the public system. While these records are subsequently screened out of the dataset, all other presentations (patients may present with multiple issues) meeting the clinical criteria for inclusion are enrolled automatically into the registry, minimising the risk of selection bias based on diagnostic or other clinical criteria.

#### Classification bias

The real-time requirement for data collection prior to the initial consultation for patients new to the practice presents challenges with respect to both information and classification bias. The registry depends on referral information to establish a minimum dataset sufficient to place a record in the relevant patient cohort (e.g. rotator cuff tear) to present the most relevant PROMs to the patient. Referral information can be highly variable in its thoroughness and accuracy, Prioritisation of patient cohorts has been used to mitigate classification bias for the cohorts of interest. However, future work may examine the potential benefits of waiting until after a consultant diagnosis has been made before sending relevant PROMs on bias or alternative screening and enrolment methods.

#### Information bias

As with any clinical registry, PRULO relies on a transformation of clinical notes and attachments into tabulated data for processing, labelling and consolidation within the registry database. While complete access to notes is available for all participating surgeon practices, the structure and level of detail of notes/observations can vary, Future work is aimed at integrating other third-party providers to automatically apply agreed coding rules on raw text, however the challenge of language harmonisation between contributors remains ongoing.

#### Data and Registry Drift

An important source of bias and measurement error in datasets retrieved from clinical registries is the potential for change over time (drift) to occur in the i) processes of the practice and ii) the processes of the registry relative to the registry dataset, as well as iii) the definitions and usage of individual variables within the registry dataset. For example, where certain clinical information has been planned to be sourced from a particular location in the electronic health record, but is now being collected by a different stakeholder in the health system and may no longer be available from the same source, or is defined differently compared to its initial usage within the registry. The PRULO registry has a core dataset and data dictionary attached and regular reporting is used to flag changes in the compliance and quality of the registry dataset and mitigate data drift.

## Data Availability

All data produced in the present work are contained in the manuscript. Registry data may be available from the investigators on request, subject to institutional approvals.

## Ethics And Dissemination

Ethical approval was obtained by the Barwon Health Research Ethics Committee (approval number 19/70). An ‘opt out’ recruitment approach was chosen for this registry. This opt-out strategy has been successfully adopted by the Australian Orthopaedic Association National Joint Research Registry (AOANJRR), and approximately 75% of Australian clinical registries (Evans et al., 2011).

## Documenting protocol amendments

Substantial changes to registry procedures will be documented via the research Protocol approved by the HREC and changes will be logged in the ANZCTR record.

## References

Colwell, C. W., Jr, Pulido, P. A., Hardwick, M. E., Sandwell, J. C., Rosen, A. S., & Copp, S. N. (2009). Initiating a multisubspecialty orthopaedic outcomes program and utilizing the data to guide practice. The Journal of Bone and Joint Surgery. American Volume, 91 Suppl 6, 134–141.

de Steiger, R. N., Miller, L. N., Davidson, D. C., Ryan, P., & Graves, S. E. (2013). Joint registry approach for identification of outlier prostheses. Acta Orthopaedica, 84(4), 348–352.

Evans, S. M., Bohensky, M., Cameron, P. A., & McNeil, J. (2011). A survey of Australian clinical registries: can quality of care be measured? Internal Medicine Journal, 41(1a), 42–48.

Gill, S., & Page, R. (2019). From big data to big impact: realizing the potential of clinical registries [Review of From big data to big impact: realizing the potential of clinical registries]. ANZ Journal of Surgery, 89(11), 1356–1357.

Gliklich, R. E., Dreyer, N. A., & Leavy, M. B. (Eds.). (2014). Registries for Evaluating Patient Outcomes: A User’s Guide (Third edition, Vol. 2). Agency for Healthcare Research and Quality, U.S. Department of Health and Human Services.

Hooper, G., Lee, A. J.-J., Rothwell, A., & Frampton, C. (2014). Current trends and projections in the utilisation rates of hip and knee replacement in New Zealand from 2001 to 2026. The New Zealand Medical Journal, 127(1401), 82–93.

Kärrholm, J. (2010). The Swedish Hip Arthroplasty Register (www.shpr.se). Acta Orthopaedica, 81(1), p3–4.

Porter, M., Armstrong, R., Howard, P., Porteous, M., & Wilkinson, J. M. (2019). Orthopaedic registries - the UK view (National Joint Registry): impact on practice. EFORT Open Reviews, 4(6), 377–390.

